# Beyond pain: Using Unsupervised Machine Learning to Identify Phenotypic Clusters of Small Fiber Neuropathy

**DOI:** 10.1101/2024.09.09.24313341

**Authors:** Peyton J. Murin, Vivian D. Gao, Stefanie Geisler

## Abstract

**Background and Objectives:** Small fiber neuropathy (SFN) is characterized by dysfunction and loss of peripheral unmyelinated and thinly myelinated nerve fibers, resulting in a phenotype that includes varying combinations of somatosensory and dysautonomia symptoms, which can be profoundly disabling and lead to decreased quality of life. Treatment aimed mainly at pain reduction, which may not target the underlying pathophysiology, is frequently ineffective. Another impediment to the effective management of SFN may be the significant between-patient heterogeneity. Accordingly, we launched this study to gain insights into the symptomatic variability of SFN and determine if SFN patients can be sub-grouped based on clinical characteristics.

**Methods:** To characterize the phenotype and investigate how patients with SFN differ from those with large fiber involvement, 105 patients with skin-biopsy proven SFN and 45 with mixed fiber neuropathy (MFN) were recruited. Using unsupervised machine learning, SFN patients were clustered based upon symptom concurrence and severity. Demographics, clinical data, symptoms, and skin biopsy- and laboratory findings were compared between the groups.

**Results:** MFN- as compared to SFN patients, were more likely to be male, older, had a lower intraepidermal nerve fiber density at the ankle and more frequent abnormal immunofixation. Beyond these differences, symptom prevalence and intensities were similar in the two cohorts. SFN patients comprised three distinct phenotypic clusters, which differed significantly in symptom severity, co-occurrence, localization, and skin biopsy findings. Only one subgroup, constituting about 20% of the patient population, was characterized by intense neuropathic pain, which was always associated with several other SFN symptoms of similarly high intensities. A pauci-symptomatic cluster comprised patients who experienced few SFN symptoms, generally of low to moderate intensity. The largest cluster was characterized by intense fatigue, myalgias and subjective weakness, but lower intensities of burning pain and paresthesia.

**Discussion:** This data-driven study introduces a new approach to subgrouping patients with SFN. Considering both neuropathic pain and pernicious symptoms beyond pain, we identified three clusters, which may be related to distinct pathophysiological mechanisms. Although additional validation will be required, our findings represent a step towards stratified treatment approaches and, ultimately, personalized treatment.

## Introduction

Small fiber neuropathy (SFN) is a neurodegenerative disease characterized by dysfunction and loss of peripheral unmyelinated C- and thinly myelinated A*δ-* fibers, which convey mechanical, chemical and thermal stimuli^1^. Both somatosensory and autonomic nerve fibers can be affected, resulting in phenotypes that include varying combinations of burning pain, tingling or prickling sensation, numbness, allodynia, muscle cramps, dry eyes or mouth, constipation, diarrhea, abnormal sweating and fatigue^2,3^. Sensory changes in SFN oftentimes manifest in a symmetric and length-dependent pattern but can also present in a non-length-dependent fashion, including focal and multifocal symptoms^4^. Although muscle strength is preserved in SFN, pain, paresthesia, and fatigue are frequently profoundly disabling and substantially decrease quality of life^5^.

Diagnostic criteria for SFN vary, but usually include a combination of typical pain symptoms, exam findings (abnormal sensation to pin and/or temperature) and decreased intraepidermal nerve fiber density (IENFD) or abnormal quantitative sensory testing (QST). Absence of large fiber signs, such as decreased sensation to vibration and proprioception and reduced muscle stretch reflexes, in combination with normal nerve conduction studies are also frequently a necessary part of the diagnosis of SFN^6^. However, inclusion/exclusion criteria vary among expert opinion guidelines and clinical studies, as do symptoms regarded as typical for SFN. While some studies consider only pain and dysautonomia as SFN symptoms^7^, others also consider non-painful manifestations, such as diminished temperature sensation^8^, while yet others also include asymptomatic patients^9^.

Many causes of SFN have been identified, including diabetes mellitus, autoimmune diseases, infections, and vitamin deficiencies^10^. However, despite extensive laboratory and genetic testing, the etiology remains unknown in more than half of the SFN patients^3^. Therapy is typically directed at the underlying cause if one can be identified. For the remainder of the patients, treatment is solely symptomatic. Although some medications may reduce pain, they may not target the underlying pathology, as mechanisms leading to SFN remain unknown in many patients.

An impediment to the effective management of SFN may be the significant between-patient heterogeneity, which reflects the manifold kinds, intensities and distributions of symptoms, etiologies, and treatment responses. This heterogeneity may be due to distinct disease mechanisms and require different treatment approaches.

Recent discoveries in rodents revealed a surprisingly complex anatomy of the small-fiber somatosensory system^11,12^. Although one characteristic of SFN is loss of intraepidermal nerve fibers in the skin, small fibers do not innervate only the skin, but also deep tissues and internal organs where axons are exposed to different microenvironments and potentially distinct toxic substances^13,14^. The somata of the somatosensory C and A-δ fibers reside in the dorsal root ganglia (DRG). With the help of advanced genetic tools, at least 15 transcriptionally distinct DRG neuron subtypes have been described of which no less than 10 subtypes give rise to C or A-δ fibers^11^. These transcriptionally identified DRG neuron populations also differ in cell body sizes, neurochemical and physiological properties, cutaneous morphologies, terminal axon branching patterns and response thresholds to different stimuli^11,15^. Many DRG neuron subtypes are further distinguished by their projections into the spinal cord, where they synapse on neurons that give rise to different ascending, parallel pathways that innervate different areas of deep brain nucle and thalamusi^16^.

Considering these recent findings, we wondered whether the complex anatomy of the sensory system contributes to the heterogeneous symptoms of patients with SFN. For instance, one could imagine that dysfunction of axons innervating the skin where they interact with keratinocytes might result in different symptoms than dysfunction of axons innervating deep tissues, such as muscles, or internal organs. Consequently, the goal of our study was two-fold. First, we sought to gain insights into the symptomatic variability in a cohort of biopsy-confirmed patients with SFN in order to expand upon information generated by the many questionnaires that focus on different pain characteristics^17,18^, whereas generally less data are available about the frequency and intensity of other kinds of SFN symptoms^19^. Secondly, we investigated whether patients can be grouped based on this information. We employed unsupervised machine learning to identify three distinct patient clusters, free from biases related to laboratory data, clinical frameworks, or patient outcomes. The clusters highlight differences in clinical symptoms and severity, skin biopsy results, and localization of symptoms, suggesting the existence of distinct phenotypes of SFN.

## Methods

### Study participants

This retrospective study was performed at the tertiary neuromuscular clinic at Washington University St. Louis. Patients presented with symptoms of SFN (including paresthesia, hyperesthesia, allodynia, fatigue, muscle cramps) and received a complete neurological examination by a board-certified Neuromuscular specialist as part of their standard clinical care. 124 of 140 patients also received EMG/NCS to evaluate for large fiber peripheral neuropathy. As part of standard clinical care, patients rated the severity of eight commonly observed SFN symptoms (i.e., burning pain, pins and needles, numbness, fatigue, muscle cramps, sore muscles, weakness, and itch)^19–23^ on a visual analog scale ranging from 0 (no symptoms) to 10 (most intense) and the frequency of seven dysautonomia symptoms (i.e., dry eyes, dry mouth, abnormal sweating, constipation or diarrhea, inability to control bowel movements, involuntary or unwanted loss of urine, and lightheadedness when getting up from a seated position) on a scale of 0 (never) to 3 (always) at time of skin biopsy. In addition, patients were asked to localize four SFN symptoms (pain, pins and needles, numbness, tingling) on a stick figure. Clinical data were extracted from medical records. Patients were diagnosed with SFN if they had abnormal pin or temperature gradient in the legs and abnormal intraepidermal nerve fiber density (IENFD) on skin biopsy^8^ and no signs of large fiber neuropathy, such as distal weakness, absent ankle reflexes, decreased sensation to vibration at the toes and/or abnormal EMG/NCS of the legs^8^. Patients were diagnosed with mixed fiber neuropathy (MFN) if they had signs of small nerve fiber dysfunction on exam, an abnormal skin biopsy, and in addition, large fiber signs on exam and/or abnormal EMG/NCS. Skin biopsies were obtained from the ankle and distal thigh using a 3.5 mm disposable punch. Tissue was processed according to standard protocols^20^ and fiber density was analyzed following well established and validated counting rules and reference values^24,25^.

### Statistical Analysis

Initial analyses comparing demographics, biopsy findings, past medical history, and laboratory findings between SFN patients and MFN patients were carried out using the chi-square test for categorical values and Mann-Whitney test for continuous variables. For the SFN patients, data were imported into Anaconda (Anaconda Software Distribution. Austin, TX). The following add-ons were used for analysis: sklearn23^26^, MatLab (The MathWorks, Inc., Natick, MA), pandas24^27^, scikit-learn-extra^26^, and sklearn.cluster^26^. The data were first framed, then the silhouette method was used to identify the ideal number of clusters as three (Supplementary Figure 1A). Thereafter, K medoids was run with a setting for three medoids. The patients were then sorted into their respective clusters, and the results were exported back to Excel. Secondary analysis was performed using Prism Version 9.5.1 (GraphPad Software, San Diego, CA). An average was calculated for each symptom’s severity for each of the clusters and an ANOVA analysis was run to assess cluster independence for each of the symptoms. Secondary analysis with Tukey’s test was used to compare the clusters to one another to confirm independence. Average symptom severity, along with a p-value, was reported for each of the symptoms. The non-parametric p-value for all comparisons was calculated using the Kruskal-Wallis test. ANOVA was used to compare the severity of each symptom in a cluster to the severity of the other symptoms in the cluster. This was followed by a post-hoc Tukey’s test. The frequency of each symptom was compared within each cluster using Kruskal-Wallis test followed by post-hoc Dunn’s multiple comparisons test. Finally, to better define localization, we compared the frequency of symptom sites using chi-square analysis, with secondary between-group comparisons for sites done using Fisher’s exact test. Statistical significance was set at p < 0.05 for all tests.

### Standard Protocol Approvals, Registrations, and Patient Consents

All procedures were approved by the human studies and ethics committee at Washington University in St. Louis.

## Results Patients

Of 145 patients included in the study (95 women, 50 men), 105 patients were diagnosed with SFN based on presence of sensory symptoms, abnormal sensation to pin and/or temperature at the feet, abnormal IENFD on skin biopsy and absence of large fiber signs (such as absent ankle reflexes, decreased muscle strength, decreased sensation to vibration, abnormal proprioception, or abnormal EMG/NCS; Fig. 1). The remaining 45 patients had symptoms and examination findings of small fiber neuropathy and an abnormal skin biopsy and, in addition, large fiber signs on exam or abnormal EMG/nerve conduction studies in the legs. These were diagnosed with mixed fiber neuropathy (MFN; Fig. 1).

**Figure 1:**
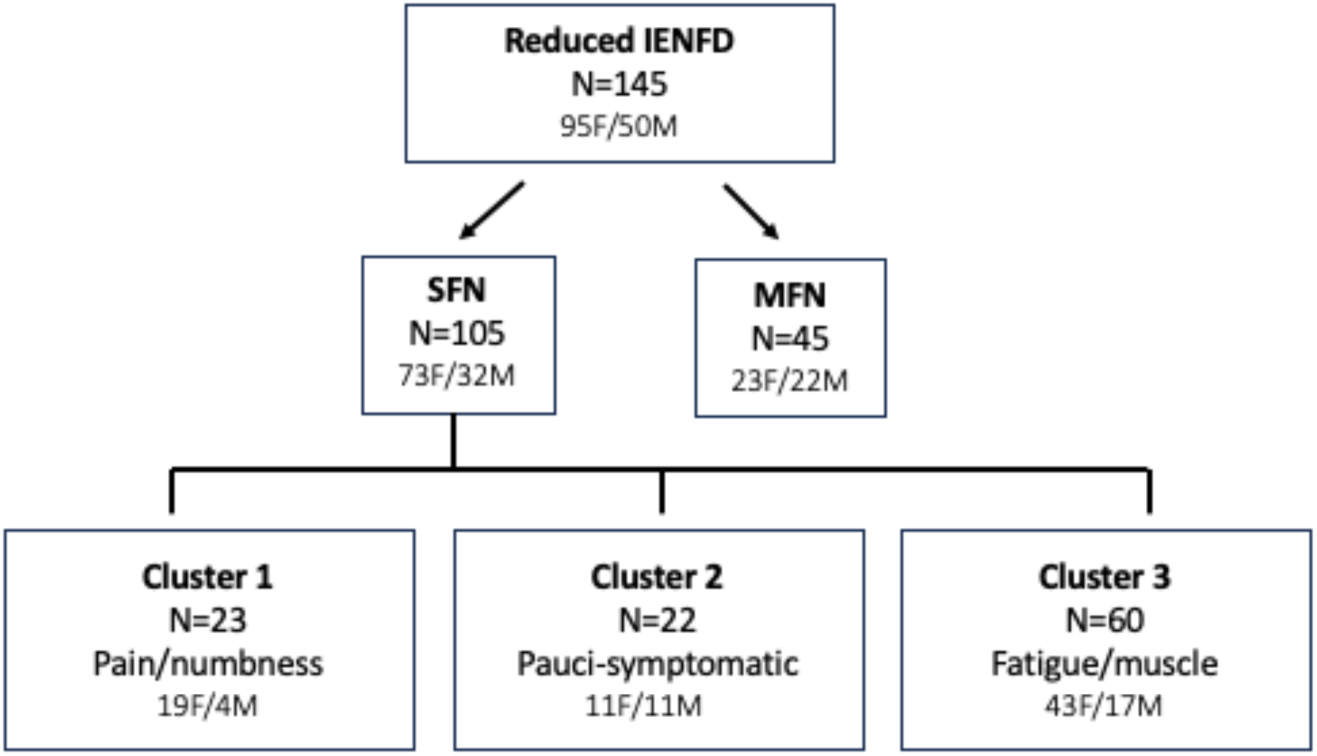
Overview of the study cohort. Of 145 patients with reduced IENFD at the ankle or thigh, 105 patients fulfilled strict criteria of SFN, which included abnormal sensation to pin and/or temperature distally, and absence of large fiber signs, such as distal weakness, absent ankle reflexes, diminished sensation to vibration, and/or abnormal EMG/NCS. 45 patients were diagnosed with MFN based on decreased IENFD, signs of SFN on exam and abnormal EMG/NCS of the legs and/or large fiber dysfunction on exam. F – female, IENFD – intra-epidermal nerve fiber density, M – male, MFN – mixed fiber neuropathy, SFN – small fiber neuropathy

## Characteristics of SFN vs MFN

The main clinical characteristics of our cohort are shown in Table 1. Patients with SFN were predominantly female (n=73, 70%) and on average 49.3±1.3 (SEM) years old. The average duration of symptoms at time of skin biopsy was 7.8±1.1 years, but varied widely, ranging from few months to several years. 21% of patients had a length-dependent loss of IENF. The most common comorbidity was hyperlipidemia (33%). In line with hyperlipidemia being frequently observed, the most frequent abnormal laboratory value was elevated lipids in 62.3% of patients. Almost half of SFN patients (46%) had positive systemic autoimmune antibodies, including positive ANA, and/or SS-A and SS-B. Although only 8.6% of SFN patients had an established diagnosis of diabetes mellitus, 37.4% of SFN patients (34/91) did have pathologically elevated HbA1c. Not all labs were tested in all patients (see Supplementary Table 1).

**Table 1:** Demographics, clinical- and laboratory findings of patients with small fiber neuropathy (SFN) and mixed fiber neuropathy (MFN). *=p<0.05, ***=p<0.001. The parametric p-value was calculated by the Chi-Square test for categorical variables. The non-parametric p-value was calculated by the Mann-Whitney test for numerical values.

|  | SFN | MFN | P value |
| --- | --- | --- | --- |
| Number of patients | 105 | 45 |  |
| Gender |  |  |  |
| • Male n (%) | 32 (30.5%) | 22 (48.9%) | **0.0313\*** |
| • Female n (%) | 73 (69.5%) | 23 (51.1%) | **0.0313\*** |
| Age at biopsy mean (range) | 49.3 (15-79) | 58.5 (29-86) | **0.0008\*\*\*** |
| Duration of symptoms, years ± SEM | 7.8 ± 1.1 | 9.0 ± 1.5 | 0.1893 |
| IENFD |  |  |  |
| • Ankle (number ± SEM) | 6.4 ± 0.4 | 5.1 ± 0.7 | **0.0234\*** |
| • Distal thigh (number ± SEM) | 9.7 ± 0.4 | 8.56 ± 0.6 | 0.1934 |
| • Intra-axonal swellings (%) | 9.5% | 11.1% | 0.7665 |
| • Length Dependent (%) | 21.0% | 33.3% | 0.1070 |
| • Non-Length Dependent (%) | 20.0% | 13.3% | 0.3301 |
| • Terminal Axonopathy (%) | 59.0% | 53.3% | 0.5167 |
| Diagnoses associated with SFN |  |  |  |
| • Diabetes mellitus (%) | 8.6% | 15.2% | 0.2220 |
| • Hyperlipidemia (%) | 33.3% | 33.3% | >0.9999 |
| • Autoimmune (%) | 10.5% | 31.1% | **0.0104\*** |
| • Fibromyalgia (%) | 10.5% | 13.3% | 0.6130 |
| • Hypothyroidism (%) | 21.0% | 11.1% | 0.1505 |
| • Alcohol abuse (%) | 1.9% | 0.0% | 0.3513 |
| • Vitamin B12 deficiency (%) | 2.9% | 4.4% | 0.6197 |
| • MGUS (%) | 2.9% | 8.9% | 0.1085 |
| Abnormal lab values |  |  |  |
| • Elevated HBA1c (%) | 37.4% | 36.8% | 0.9555 |
| • Elevated lipids (%) | 62.3% | 75.0% | 0.2036 |
| • Autoimmune systemic (%) | 45.6% | 36.6% | 0.3359 |
| • Autoimmune nerve (%) | 31.1% | 32.0% | 0.9317 |
| • Elevated TSH (%) | 6.3% | 9.8% | 0.4700 |
| • Low Vitamin B12 (%) | 0% | 2.3% | 0.1378 |
| • Abnormal immunofixation/SPEP (%) | 5.8% | 18.4% | **0.0269\*** |

MFN patients were more likely to be male (48.9% vs 30.5%, p = 0.031, Table 1) and were on average older at the time of diagnosis (58.5 vs 49.3 years; p = 0.001, Table 1). Nevertheless, symptom duration before skin biopsy was similar between MFN and SFN patients (7.8 years SFN, 9.0 years MFN; p = 0.1893). Patients with MFN had a significantly lower IENFD at the ankle (5.1±0.7 axons/mm) when compared to patients with SFN (6.4±0.4 axons/mm, p=0.023, Mann Whitney U test). Rates of medical comorbidities were similar between SFN and MFN patients, except for overall autoimmune diseases, which were diagnosed more frequently in MFN patients (31.1% vs 10.5%, p = 0.010). The only abnormal laboratory test that had a significantly different prevalence was abnormal immunofixation, which was more commonly observed in MFN as compared to SFN patients (18.4% vs 5.8%, p = 0.027).

The most prevalent and intense symptom in both cohorts was fatigue, which was reported by 77.1% of SFN- and 82.2% of MFN patients with an average severity of 5/10 in both cohorts (Fig. 2). While previous studies showed higher intensity pain in large fiber- as compared to small fiber neuropathy patients, comparisons between the MFN and SFN cohorts yielded no differences in symptoms frequency or severity. The symptom of itch was significantly less frequently observed in patients of both SFN and MFN cohorts and was significantly less severe than the remaining symptoms (Fig. 2).

**Figure 2:**
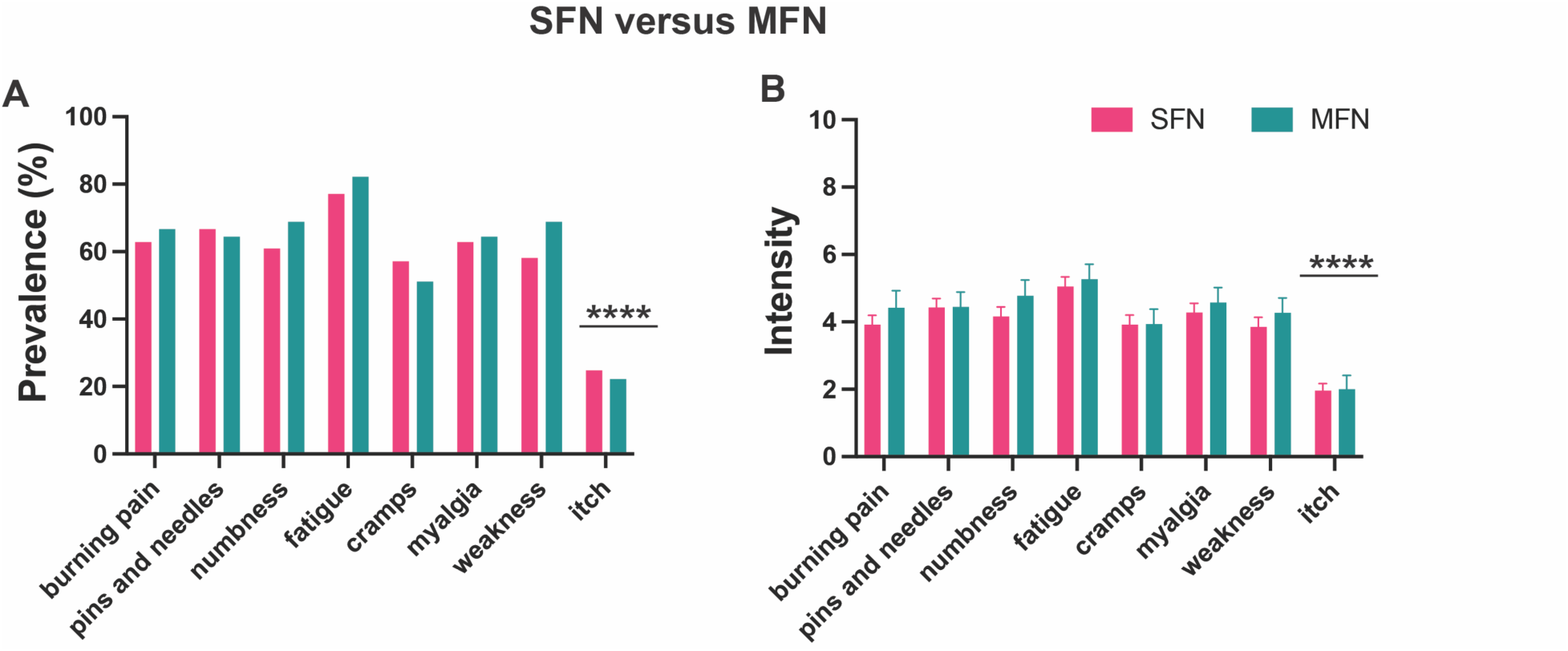
Symptom prevalence and severity of patients with small fiber neuropathy (SFN) and mixed fiber neuropathy (MFN). The prevalence (A) and severity (B) of symptoms was similar between patients with SFN and MFN. The symptom of ‘itch’ was reported by significantly fewer patients (A) and of significantly less intensity (B) by both patients with SFN (p < 0.0001, Kruskal-Wallis) and MFN (p < 0.0001).

Thus, as a population, SFN patients were more likely to be female and younger than MFN patients. MFN patients had a lesser IENFD at the ankle and were more likely to have abnormal immunofixation and comorbid autoimmune disease than SFN patients (Table 1). Beyond large fiber degeneration, reduced ankle IENFD, autoimmune labs and immunofixation, the symptoms experienced by patients with MFN and SFN were similar.

## SFN phenotypes

### Symptom intensity

To determine whether certain SFN symptoms group together and thus may represent distinct phenotypes, SFN patients were asked to rate the intensity of symptoms they experienced during the past week on a VAS (see Methods). The silhouette method was used to identify the ideal number of clusters as three (Fig. S1A). Next, K medoids was run with a setting for three medoids, and the patients were sorted into their respective clusters: cluster 1 consisting of 23 patients, cluster 2 of 22 patients and cluster 3 of 60 patients. To determine if the clusters were distinct from one another, ANOVA analysis was used to compare the symptom severity and dysautonomia symptom frequency between the clusters. This analysis resulted in statistically significant variance for seven of eight SFN symptoms (Fig. 1SB) and all dysautonomia symptoms (Fig S1C). To confirm the independence of the clusters, Tukey’s test was used, which again identified statistically significant differences for seven of the eight SFN symptoms. The symptom of “itch” was not significantly different between the three clusters by ANOVA analysis or in the follow-up Tukey’s test comparisons.

Next, we sought to identify defining characteristics of the clusters. Symptoms were matched, and a color-coded heat map was created (Fig. 3). Patients in cluster 1 suffered from many symptoms at high intensities (Fig. 4A) with an average of 7.5/10 for “numbness” and 7.2/10 for “burning pain” (Fig. 4A). Fatigue (average 7.2/10) and muscle cramps (average 7.1/10) were also experienced as severe, whereas subjective weakness had a slightly lower intensity of 5.8/10. Itch was significantly lower than all other symptoms at an intensity of 2.8. Thus, because intense burning pain was accompanied by similarly intense numbness in this subgroup (Fig. 3, 4A), we call this cluster “Pain/numbness predominant.”

**Figure 3:**
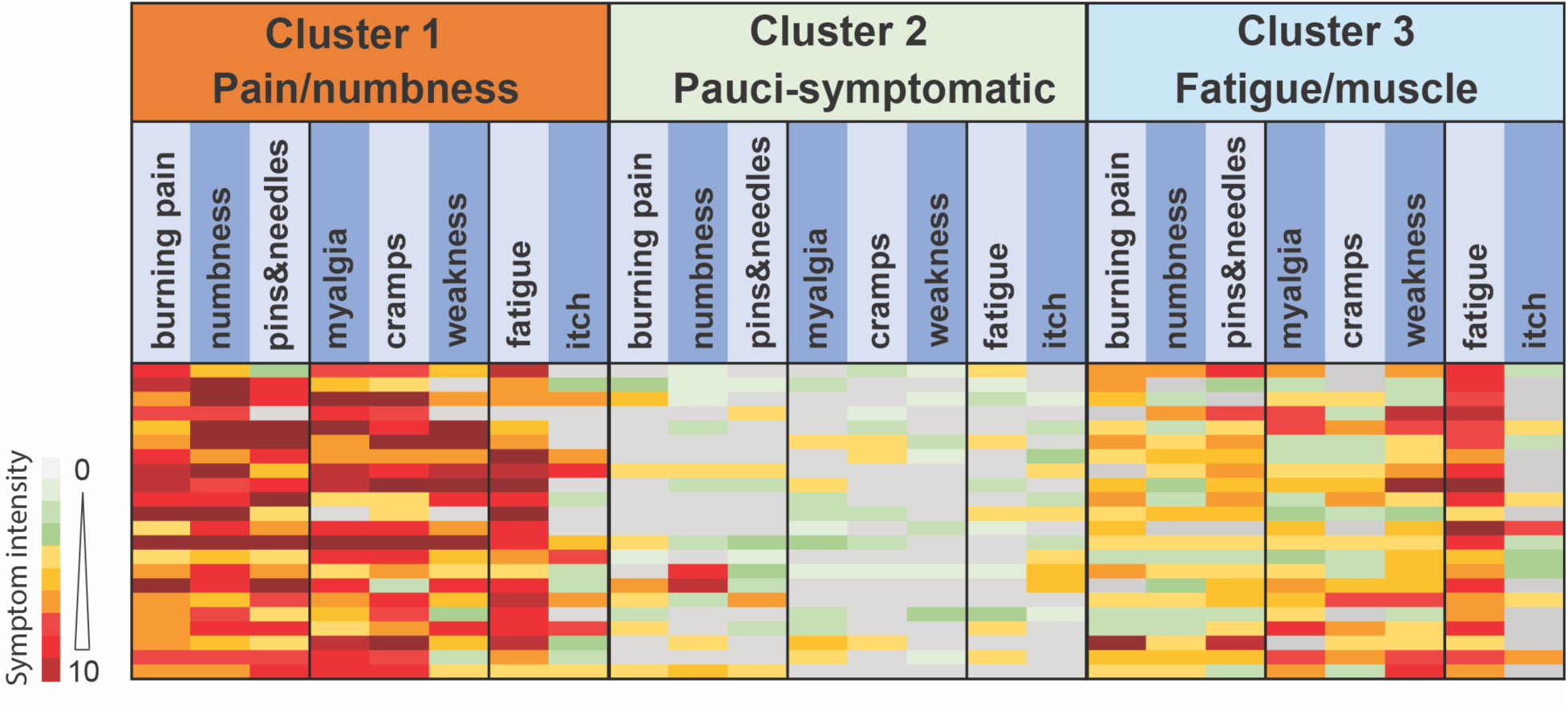
Heatmap of SFN symptom severities of twenty-two representative patients of each cluster. Patients marked the severity of each symptom they experienced during the past week on a visual analog scale ranging from 0 (no symptoms) to 10 (worst possible). Each row in each cluster represents one individual patient. Note the high severity of most symptoms in cluster 1 (pain/numbness) and the high severity of fatigue in the cluster 3 (fatigue/muscle predominant), whereas patients in cluster 2 experienced mostly symptoms of low to moderate severities.

**Figure 4:**
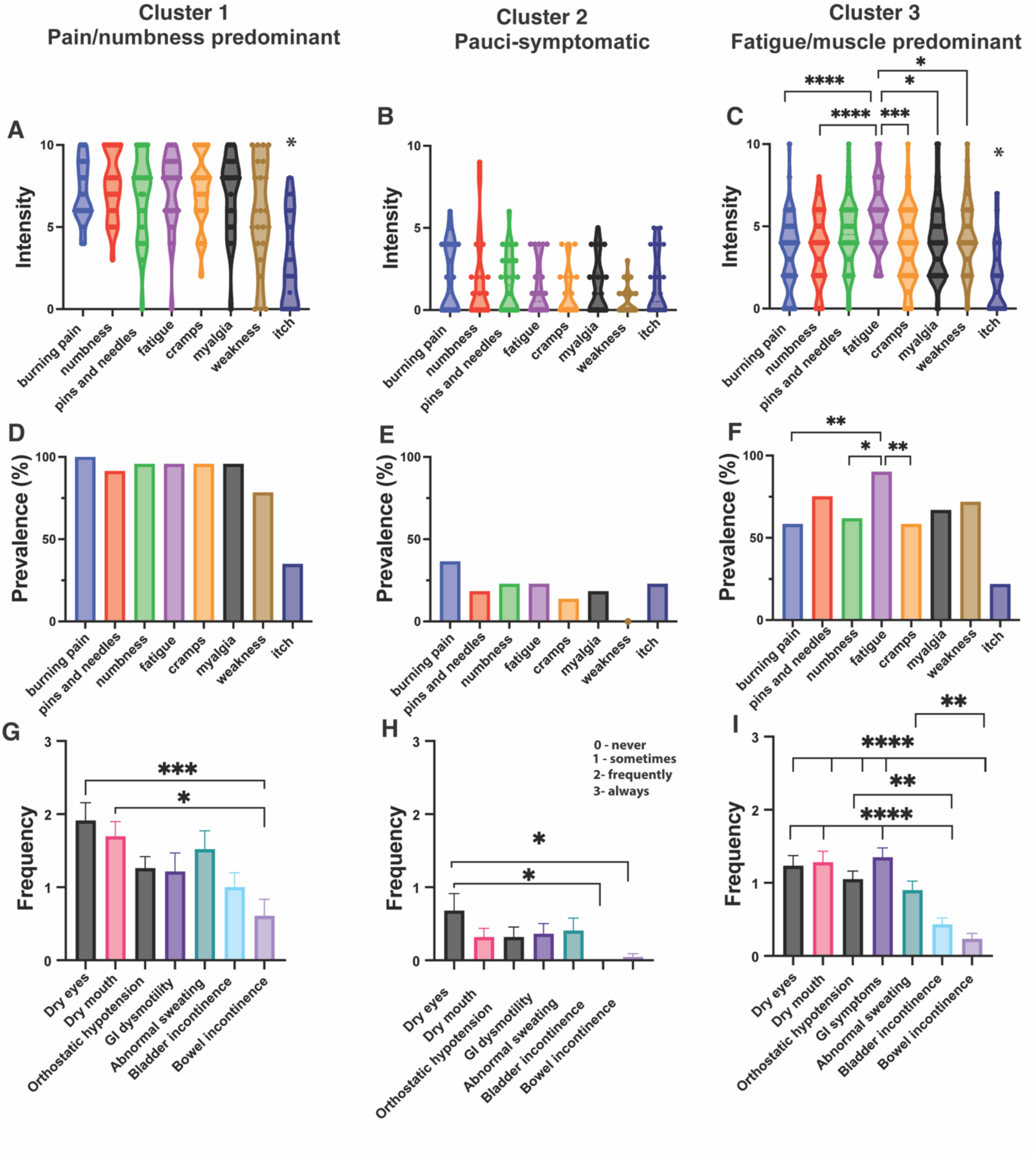
Symptom severity and prevalence of SFN patients. **A-C** Comparison of symptom intensity in each cluster. **A**: Patients in cluster 1 experienced several symptoms at high intensity with the highest severity for numbness and burning pain. *Itch was of significantly less intensity than the other symptoms (p < 0.0001, one-way ANOVA). **B**: Patients in cluster 2 experienced symptoms as generally less severe. **C**: Patients in cluster 3 had significantly more severe fatigue than burning pain (p < 0.0001, one-way ANOVA), numbness (p < 0.0001), cramps (p = 0.0001), myalgia (p = 0.0195), weakness (p = 0.0294) and itch (p = 0.0011). **D-F**: Prevalence of each symptom in the three clusters. The symptom of fatigue was reported by significantly more patients than burning pain (p = 0.0094, Kruskal-Wallis), numbness (p = 0.0372) and muscle cramps (p = 0.0094). **G-H** Patients ranked the frequency for each dysautonomia symptom during the past week ranging from never (0) to always (3). **G**: Patients in cluster 1 had significantly less bowel incontinence than dry eyes (p = 0.009, one-way ANOVA) and dry mouth (p = 0.0112). **H:** Patients in cluster 2 had significantly less bowel (p = 0.0269, one-way ANOVA) and bladder incontinence (p = 0.0133) than dry eyes. **I:** Patients in cluster 3 experienced significantly less bowel and bladder incontinence than all other dysautonomia symptoms (p < 0.0001, one-way ANOVA). * = p < 0.05, ** = p <0.01, *** =p < 0.001 and **** = p < 0.0001.

Patients in cluster 2 expressed few symptoms, all of low intensity (Fig. 3), with the highest average intensity of 2.0 for burning pain and the lowest of 0.5 for weakness and muscle cramps (Fig. 4B), leading us to call this cluster “Pauci-symptomatic.” The third cluster (n = 60) was characterized by patients who exhibited mostly symptoms of moderate severity with one or two symptoms of higher intensity (Fig. 3). Upon further inspection, patients in cluster 3 had especially severe fatigue (average 5.6; Figs. 3, 4C), which was significantly more intense than intensities for burning pain, numbness, cramps, myalgia, subjective weakness, and itch (Fig. 4C). In contrast to cluster 1, in which patients had especially high intensities of burning pain and numbness and lower intensity of weakness, patients in cluster 3 had more severe subjective weakness and myalgia (average 4.3) than burning pain/numbness (average 3.6; t-test p=0.0085). Therefore, we called cluster 3 “Fatigue/muscle predominant.”

### Symptom prevalence

Next, we sought to determine whether certain symptoms occur together more commonly than others. To do so, we focused on symptoms with moderate to severe intensity and converted symptoms with a severity of 0-3 to a value of ‘0’ and symptoms with a severity of 4-10 to a value of ‘1’ and used the Kruskal-Wallis and Dunn’s multiple comparison test for statistical analysis. In the pain/numbness cluster, most symptoms were reported by 90 to 100% of patients (Fig. 4D), whereas subjective weakness was noted by only 78% and itch by 35%. In the pauci-symptomatic cluster, 36% of patients experienced burning pain, whereas nobody noticed subjective weakness (Fig. 4E). In the fatigue/muscle predominant cluster, fatigue was reported by significantly more patients (90%) than burning pain (58%), numbness (61%), and muscle cramps (58%; Fig. 4F), whereas myalgia and weakness were named by 71% and 67% of patients.

Thus, in cluster 1 (pain/numbness) most patients experienced several symptoms at high intensities, in cluster 2 (pauci-symptomatic) patients exhibited only few symptoms and in cluster 3 (fatigue/muscle predominant) the greatest percentage of patients had fatigue, which was also of highest intensity, followed by symptoms of myalgia and subjective weakness.

### Symptom localization

The localization data for burning pain, pins and needles, numbness and tingling can be seen in Supplementary Table 2 and Figure 5. Despite differences in symptom severity, we expected similar anatomical distributions of symptoms. To assess this, we reported a frequency for each anatomical site marked on the stick figures and used chi-square and Fisher’s exact test for comparisons. Interestingly, burning pain, pins and needles, numbness and tingling were not generally observed in the same location in individual patients, and this was also reflected at the group level by different patterns of symptom distribution within clusters (Fig 5). Consistent with the oft-reported observation that SFN symptoms start in the feet, most patients reported symptoms in feet in all clusters. However, the involvement of more proximal sites differed between groups (Fig 5).

**Figure 5:**
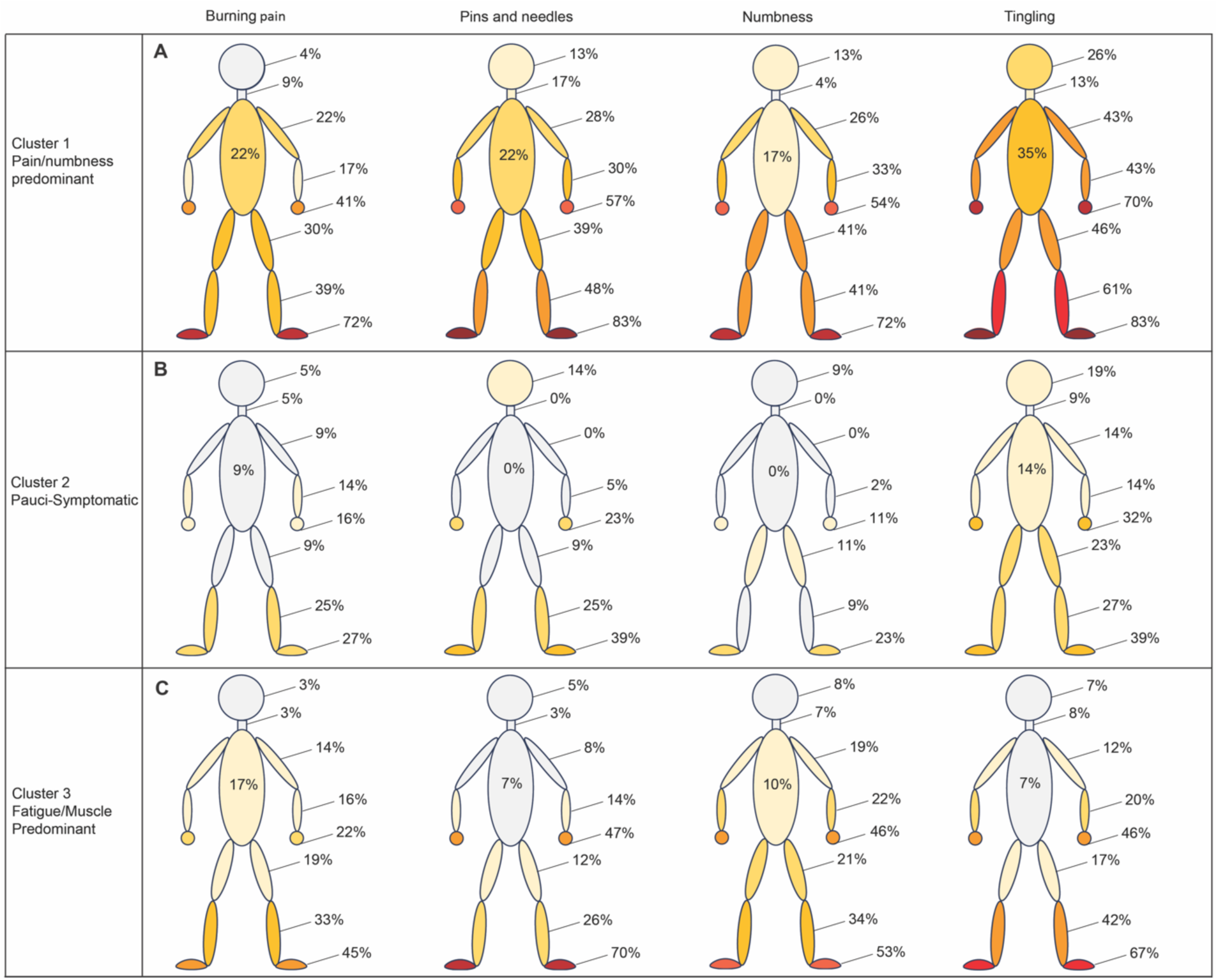
Localization of four SFN symptoms in each cluster. Shown is the percent of patients in each cluster who marked the location of the corresponding symptom on a stick-figure.

Patients in the pain/numbness predominant cluster had the broadest distribution of symptoms, largely in a length-dependent pattern (Fig. 5A). In this cluster, the most frequently marked symptom of tingling was reported in feet and hands by 83% and 70% of patients, respectively, whereas upper legs and upper arms were involved in 46% and 43% of patients. 35% of patients observed tingling in the torso and 26% in the head. Burning pain, pins and needles and numbness followed a similar length-dependent pattern in this cluster. Although burning pain was most intense and most broadly distributed in the pain/numbness cluster, only feet and hands were significantly more frequently affected when compared to the other clusters, suggesting that burning pain is relatively more common in the proximal areas in clusters 2 and 3. Furthermore, 83% of patients in the pain/numbness cluster and only 39% of those in the pauci-symptomatic cluster (Fig. 5B) marked pins and needles in the feet, whereas the prevalence of pins and needles in the head area was similar between the two clusters, with 14% of patients in the pauci-symptomatic group and 13% in the pain/numbness group reporting this symptom (Fig. 5A, B). Thus, in patients with the overall highest intensity of symptoms (pain/numbness group), symptoms also were most widely distributed and followed a length-dependent pattern. Symptoms were more localized and involved more proximal sites in the other two clusters (Fig 5).

### Dysautonomia

Symptoms of dysautonomia exhibited the same patterns as somatosensory symptoms in the sense that patients of the pain/numbness cluster most frequently reported dysautonomia symptoms, except GI symptoms, which were noted with similar frequencies by patients in the pain/numbness and fatigue/muscle clusters (Fig. 4G-I). The most frequently occurring dysautonomia symptoms in patients of the pain/numbness cluster were dry eyes, dry mouth and abnormal sweating, all of which occurred frequently in that group during the week prior to assessment (Fig. 4G). Patients in the fatigue/muscle cluster noted most frequently GI symptoms, followed by dry eyes and dry mouth (Fig. 4I). Bowel incontinence was observed significantly less often in all clusters as compared to all other symptoms of dysautonomia.

### Cluster demographics and comorbidities

The main clinical characteristics of the clusters are described in Table 2. The patients in the pain/numbness predominant cluster tended to be older (53.4 years vs 49.8 years, fatigue/muscle vs 43.8 years pauci-symptomatic) and predominantly female (82.6% vs 71.7% fatigue/muscle vs 50.0% pauci-symptomatic), with a longer duration of symptoms prior to biopsy (14.7 years vs 6.2 years fatigue/muscle vs 5.1 years pauci-symptomatic), however these differences were not statistically significant (Table 2).

**Table 2:**
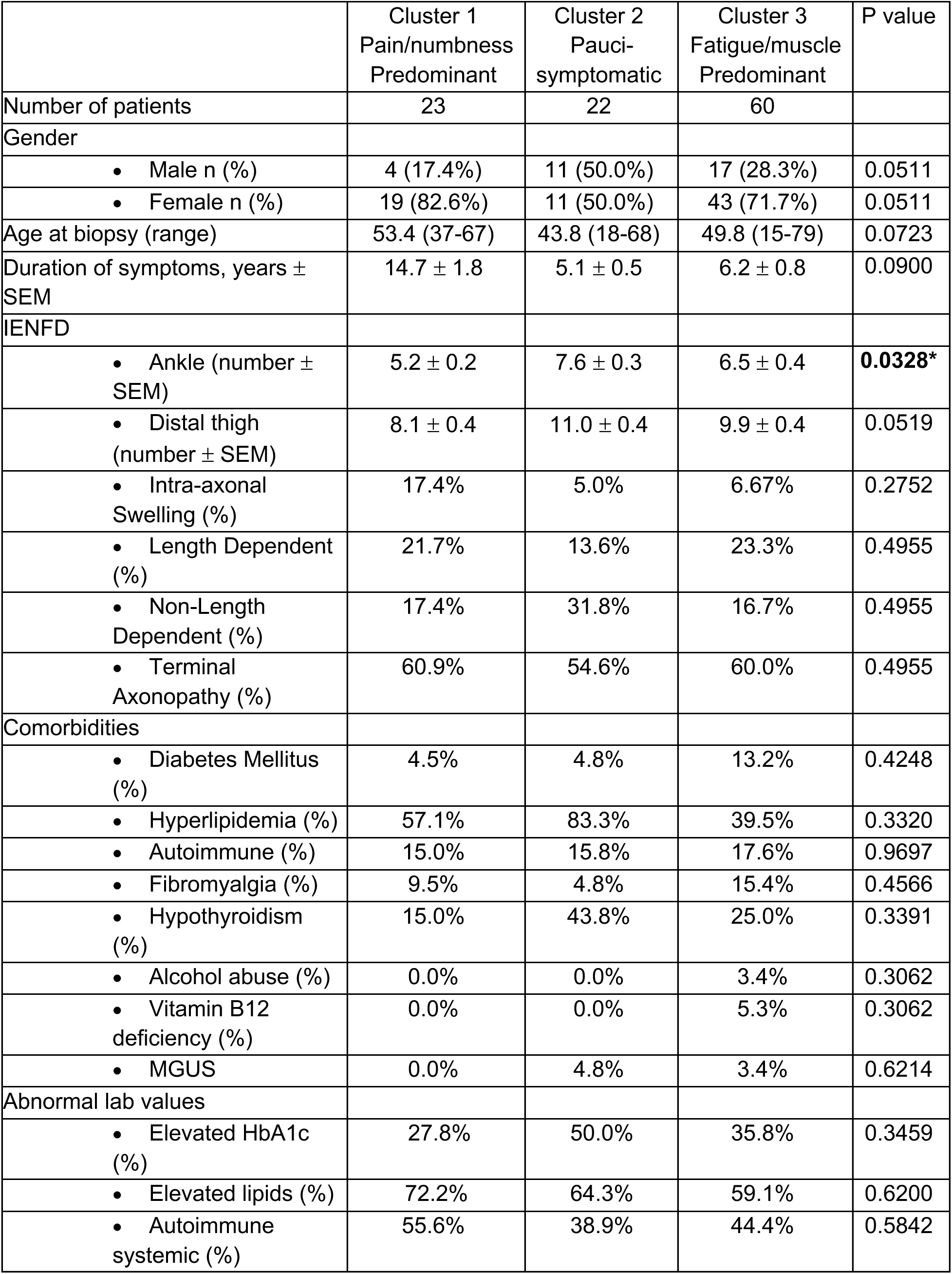
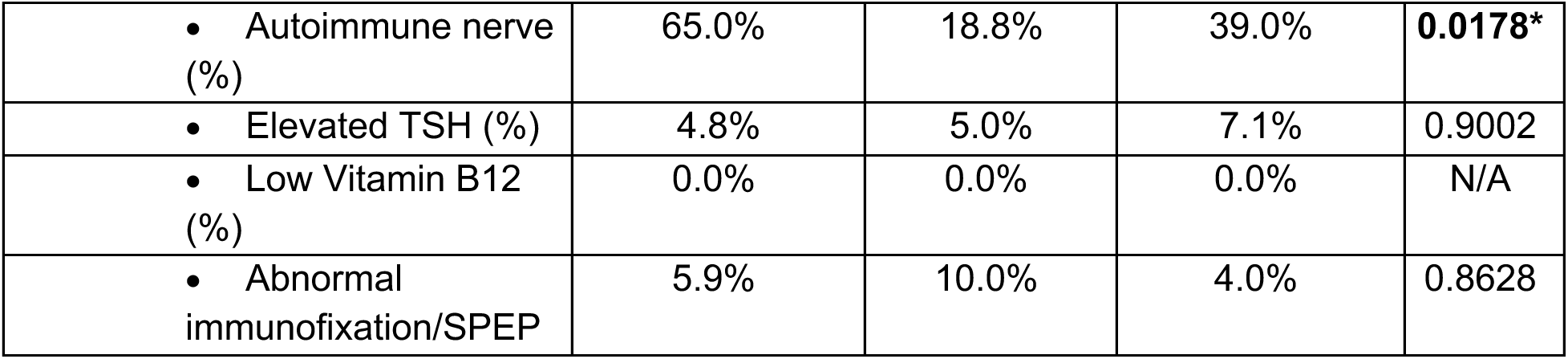
Demographics, clinical, and laboratory findings between the SFN phenotypic clusters. Length-dependent loss indicates preferential IENFD at the ankle than thigh (IENFD ankle/thigh ratio > 2), non-length dependent loss indicates preferentially loss at the thigh (ankle/thigh ratio < 1) and terminal axonopathy indicates similar loss at the ankle and thigh^50^. Autoimmune systemic includes antibodies to ANA, ANCA and ENA; autoimmune nerve includes antibodies to TS-HDS, FGFR3, and plexin; MGUS – monoclonal gammopathy of unknown significance. The parametric p-value was calculated by the Chi-Square test for categorical variables and the non-parametric p-value by the Kruskal-Wallis test for numerical values. *=p< 0.05

|  | Cluster 1<br>Pain/numbness<br>Predominant | Cluster 2<br>Pauci-symptomatic | Cluster 3<br>Fatigue/muscle<br>Predominant | P value |
| --- | --- | --- | --- | --- |
| Number of patients | 23 | 22 | 60 |  |
| Gender |  |  |  |  |
| • Male n (%) | 4 (17.4%) | 11 (50.0%) | 17 (28.3%) | 0.0511 |
| • Female n (%) | 19 (82.6%) | 11 (50.0%) | 43 (71.7%) | 0.0511 |
| Age at biopsy (range) | 53.4 (37-67) | 43.8 (18-68) | 49.8 (15-79) | 0.0723 |
| Duration of symptoms, years ± SEM | 14.7 ± 1.8 | 5.1 ± 0.5 | 6.2 ± 0.8 | 0.0900 |
| IENFD |  |  |  |  |
| • Ankle (number ± SEM) | 5.2 ± 0.2 | 7.6 ± 0.3 | 6.5 ± 0.4 | **0.0328\*** |
| • Distal thigh (number ± SEM) | 8.1 ± 0.4 | 11.0 ± 0.4 | 9.9 ± 0.4 | 0.0519 |
| • Intra-axonal Swelling (%) | 17.4% | 5.0% | 6.67% | 0.2752 |
| • Length Dependent (%) | 21.7% | 13.6% | 23.3% | 0.4955 |
| • Non-Length Dependent (%) | 17.4% | 31.8% | 16.7% | 0.4955 |
| • Terminal Axonopathy (%) | 60.9% | 54.6% | 60.0% | 0.4955 |
| Comorbidities |  |  |  |  |
| • Diabetes Mellitus (%) | 4.5% | 4.8% | 13.2% | 0.4248 |
| • Hyperlipidemia (%) | 57.1% | 83.3% | 39.5% | 0.3320 |
| • Autoimmune (%) | 15.0% | 15.8% | 17.6% | 0.9697 |
| • Fibromyalgia (%) | 9.5% | 4.8% | 15.4% | 0.4566 |
| • Hypothyroidism (%) | 15.0% | 43.8% | 25.0% | 0.3391 |
| • Alcohol abuse (%) | 0.0% | 0.0% | 3.4% | 0.3062 |
| • Vitamin B12 deficiency (%) | 0.0% | 0.0% | 5.3% | 0.3062 |
| • MGUS | 0.0% | 4.8% | 3.4% | 0.6214 |
| Abnormal lab values |  |  |  |  |
| • Elevated HbA1c (%) | 27.8% | 50.0% | 35.8% | 0.3459 |
| • Elevated lipids (%) | 72.2% | 64.3% | 59.1% | 0.6200 |
| • Autoimmune systemic (%) | 55.6% | 38.9% | 44.4% | 0.5842 |
| • Autoimmune nerve (%) | 65.0% | 18.8% | 39.0% | **0.0178\*** |
| • Elevated TSH (%) | 4.8% | 5.0% | 7.1% | 0.9002 |
| • Low Vitamin B12 (%) | 0.0% | 0.0% | 0.0% | N/A |
| • Abnormal immunofixation/SPEP | 5.9% | 10.0% | 4.0% | 0.8628 |

While previous literature has not supported an association between IENFD and pain^8^, patients in the pain/numbness predominant cluster had significantly lower IENFD at the ankle, suggesting that considering several SFN symptoms in aggregate may more precisely reflect disease. There were no differences in the form of intra-axonal swellings or the pattern of axon degeneration, with 14% to 23% of patients showing a length-dependent loss of intraepidermal axons across the clusters. Interestingly, the one lab value that differed significantly between clusters was autoantibodies against nerves, including TS-HDS, FGFR3 and plexin, which was found in 65% of tested patients in the pain/numbness cluster, but only 39% and 19% of tested patients in the pauci-symptomatic and fatigue/muscle clusters, respectively. Thus, the patients in the pain/numbness cluster had significantly reduced IENFD at the ankle, and significantly more abnormal anti-nerve antibodies than the other clusters.

## Discussion

SFN typically has been defined by its neuropathic pain symptoms, including burning pain and allodynia^28^, but less is known about the prevalence, intensity and co-occurrence of other symptoms^19^. Here, we characterized a large, well-defined and skin biopsy confirmed cohort of SFN patients among which we identified three distinct SFN symptom clusters: ‘pain/numbness predominant,’ ‘pauci-symptomatic,’ and ‘fatigue/muscle predominant.’ The clusters are distinct in terms of severity and co-occurrence, skin biopsy findings and antibodies, suggesting the clusters represent distinct SFN phenotypes.

Focusing on eight SFN symptoms, we find that in only one subgroup, comprising about 20% of the patient population, was neuropathic pain the most intense and prevalent symptom and always associated with several other high intensity SFN symptoms, including numbness (Fig. 3). As numbness is frequently the result of axon degeneration^29^, our findings suggest that intense pain in SFN is commonly accompanied by axon loss. Therefore, in addition to targeting nociceptor ion channels^30^, blocking axon degeneration, for instance with novel SARM1 inhibitors^31^, may be a promising therapeutic avenue in SFN patients with intense neuropathic pain.

The biggest cluster in our cohort was characterized by fatigue, which was more severe and prevalent than the other symptoms in the cluster. This is in accord with a previous study that also revealed fatigue as one of five most prevalent symptoms in SFN^19^. Fatigue is a well described symptom in several neurological disorders, including multiple sclerosis^32^, stroke^33^, Parkinson’s Disease and other movement disorders^34^, but is rarely assessed in standard neuropathic pain screening questionnaires^35^. Recent work identified associations between chronic fatigue syndrome and SFN in post-acute COVID-19 patients^36^ and between reduced IENFD and fatigue in complex regional pain syndrome^37^. Here, we extend this finding by suggesting that fatigue is a common, high intensity symptom of SFN for which treatments should be developed. The pathophysiology of fatigue has been hypothesized to be secondary to an overactive or dysregulated immune response^38^ resulting in both higher and lower levels of pro-inflammatory cytokines^39^. Given the more generalized localization of symptoms and the high intensity of fatigue in most patients in cluster 3, it is tempting to speculate that an immune dysfunction contributes to the symptom presentation in that cluster.

The pauci-symptomatic cluster was characterized by patients who experienced only a few symptoms of low to moderate intensities. While the other two clusters are female predominant (83%/17% and 72%/28% F/M), this one involves more males (50%/50% F/M). It is possible that patients in this cluster are earlier in the disease course, and that male patients progress slower than females (as there are fewer males in the more symptom-intense clusters). However, the duration of symptoms in the pauci-symptomatic cluster was similar to the fatigue/muscle cluster (5 vs 6 years), suggesting that additional mechanisms are involved. RNA sequencing of human DRGs from patients suffering from pain revealed major sex differences in the gene expression pattern between males and females^40^. Similarly, male and female patients with chronic fatigue syndrome express different subpopulations of immune cells and distinct subsets of differentially expressed genes, suggesting that the biological processes underlying fatigue in male and female patients^41^ are distinct. Accordingly, sex-specific differences in DRG gene expression patterns and neuro-immune functions may contribute to the less severe phenotype that characterizes this cluster.

Itch has been reported as a symptom of SFN, possibly due to damage of epidermal C-fibers^42^. Pathophysiology of itch involves activation of epidermal and dermal C-fibers by pruritogens. Studies in mice suggest that these C-fibers transmit itch exclusively^43^. Itch was reported with significantly lower intensity and by fewer patients than the other SFN symptoms in our cohort. Separate itch-specific C-fibers, transmission pathways, and dorsal root ganglion neurons^16^ provide a possible explanation for lesser itch frequency and severity compared to the overall SFN phenotype. Why itch-selective units are less vulnerable to neuropathic processes will be an interesting avenue for future research.

Prior literature comparing SFN and MFN is limited^44^. Our study provides a comprehensive demographic, clinical, and histopathological comparison of patients meeting strict criteria of SFN and MFN. We find that SFN patients were younger and more likely to be female than patients with MFN. While more intense pain was reported in SFN patients compared to patients with large fiber neuropathy^8^, we did not observe a difference in symptom frequency or intensity, including pain, between SFN and MFN patients, possibly because MFN patients in our cohort likely had more small fiber involvement than the large fiber neuropathy patients in the previous study. Notably, our MFN patients had significantly more loss of IENFD than SFN patients. The combination of older age and lesser IENFD on skin biopsy could suggest progression of the disease, and indeed, progression from SFN to MFN has been demonstrated in diabetic neuropathy^28^. However, there was no difference in the time from symptom onset to skin biopsy in our cohorts. Importantly, just as SFN may progress into MFN, there are reports of the large fiber neuropathy progressing into MFN^44^, suggesting that any pathophysiological progression from one to the other is not necessarily linear or directional.

While several subtyping and clustering approaches have been used to discover pain-associated phenotypes^45–47^, subtyping of SFN is limited^2,19^. Although clustering based on pain phenotypes can extend to patients with peripheral neuropathy, those subgroupings typically have included mixes of patients with diverse pain conditions, including back pain and herpetic neuralgia^48^, and likely are not generalizable to patients with SFN. Furthermore, as demonstrated here and previously, not all patients with SFN experience pain^19^, which thus excludes a sizable population of SFN patients if pain is a mandatory inclusion criterion.

Our approach offers several strengths. First, we were able to effectively cluster all study participants, whereas many of the previous studies left a significant number of patients as ‘idiopathic’ or without a subgroup. Second, our approach demonstrated robust independence in both symptom severity and co-occurrence. Third, our phenotypes displayed a difference in IENFD whereas previous work has not demonstrated a correlation between neuropathic pain and IENFD in SFN^49^, possibly because we considered the entire phenotypes including less commonly considered SFN symptoms, such as fatigue and cramps. A further advantage of our approach is the ease of adaptability to the clinical setting. The study survey consists only of a simple, 15-symptom clinical questionnaire, making it practical for busy clinicians. The large number of well-characterized, biopsy-confirmed SFN patients (n = 105), the strict inclusion criteria, the collection of data by a single investigator, and the utilization of machine learning are further strengths. These phenotypic clusters may reflect different pathophysiological mechanisms resulting from select involvement of certain DRG neuron subpopulations. Alternatively the clusters could reflect different subjective experiences of the same pathological processes. To distinguish between these alternatives and possibly others will require further investigation.

## Conclusions

Here we provide a careful description of a cohort of biopsy-proven SFN patients. Using unsupervised machine learning, we have identified three phenotypic clusters of SFN, which differ in symptom severity and concurrence and dysautonomia frequency. Importantly, by considering entire phenotypes, we were able to identify an association between clinical phenotype and skin biopsy findings, suggesting the clinical differences represent distinct pathophysiologies. In doing so, we highlight the key role of fatigue as a symptom and emphasize the importance of considering symptoms beyond neuropathic pain in the assessment and treatment of SFN patients.

## Data Availability

All data produced in the present study are available upon reasonable request to the authors

## Acknowledgement

The study was supported by National Institutes of Health grant R37CA267905, the NIH/National Center for Advancing Translational Sciences (NCATS), grant UL1 TR002345 and the Foundation for Barnes-Jewish Hospital.

---

**Supplementary Table 1:** Percent of patients who received the indicated clinical test in each group.

| **Lab Value** | **SFN**<br>**(n =**<br>**105)** | **MFN**<br>**(n = 45)** | **Pain/numbness**<br>**predominant**<br>**(n = 22)** | **Pauci-**<br>**symptomatic**<br>**(n = 23)** | **Fatigue/muscle**<br>**predominant**<br>**(n = 60)** |
| --- | --- | --- | --- | --- | --- |
| Skin Biopsy | 100% | 100% | 100% | 100% | 100% |
| Vit B12 | 89.5% | 95.6% | 86.4% | 87.0% | 91.7% |
| TSH | 100% | 100% | 100% | 100% | 100% |
| MGUS | 100% | 100% | 100% | 100% | 100% |
| HbA1c | 86.7% | 84.4% | 81.8% | 87.0% | 83.3% |
| Lipids | 73.3% | 71.1% | 81.8% | 60.9% | 73.3% |
| Autoimmune –<br>Systemic | 85.7% | 91.1% | 81.8% | 78.3% | 90.0% |
| Autoimmune –<br>Nerve | 70.5% | 55.6% | 90.9% | 69.6% | 68.4% |
| Abnormal<br>Immunofixation | 82.9% | 84.4% | 77.3% | 87.0% | 83.3% |

**Supplementary Table 2:** Percent of patients in each SFN cluster who marked the indicated symptom on a stick figure. Chi-Square test.

|  | Pain/numbness<br>Predominant | Pauci-symptomatic | Fatigue/muscle<br>Predominant | P value |
| --- | --- | --- | --- | --- |
| Number of patients | 23 | 22 | 60 |  |
| Burning Pain |  |  |  |  |
| • Feet (%) | 72.0% | 27.0% | 45.0% | **0.0006\*** |
| • Lower Legs (%) | 39.0 | 25.0% | 33.0% | 0.4639 |
| • Upper Legs (%) | 30.0% | 9.0% | 19.0% | 0.1929 |
| • Hands (%) | 41.0% | 16.0% | 22.0% | **0.0142\*** |
| • Lower Arms (%) | 17.0% | 14.0% | 16.0% | 0.9941 |
| • Upper Arms (%) | 22.0% | 9.0% | 14.0% | 0.4728 |
| • Torso (%) | 22.0% | 9.0% | 17.0% | 0.5094 |
| • Head (%) | 4.0% | 5.0% | 3.0% | 0.9508 |
| • Neck (%) | 9.0% | 5.0% | 3.0% | 0.5895 |
| Pins and Needles |  |  |  |  |
| • Feet (%) | 83.0% | 39.0% | 70.0% | **0.0002\*** |
| • Lower Legs (%) | 48.0% | 25.0% | 26.0% | **0.0857** |
| • Upper Legs (%) | 39.0% | 9.0% | 12.0% | **0.0064\*** |
| • Hands (%) | 57.0% | 23.0% | 47.0% | 0.0584 |
| • Lower Arms (%) | 30.0% | 5.0% | 14.0% | **0.0444\*** |
| • Upper Arms (%) | 28.0% | 0.0% | 8.0% | **0.0016\*** |
| • Torso (%) | 22.0% | 0.0% | 7.0% | **0.0244\*** |
| • Head (%) | 13.0% | 14.0% | 5.0% | 0.3192 |
| • Neck (%) | 17.0% | 0.0 | 3.0% | **0.0204\*** |
| Numbness |  |  |  |  |
| • Feet (%) | 72.0% | 23.0% | 53.0% | **0.0007\*** |
| • Lower Legs (%) | 41.0% | 9.0% | 34.0% | **0.0267\*** |
| • Upper Legs (%) | 41.0% | 11.0% | 21.0% | **0.0022\*** |
| • Hands (%) | 54.0% | 11.0% | 46.0% | **<0.0001\*** |
| • Lower Arms (%) | 33.0% | 2.0% | 22.0% | **0.0011\*** |
| • Upper Arms (%) | 26.0% | 0.0% | 19.0% | **0.0470\*** |
| • Torso (%) | 17.0% | 0.0% | 10.0% | 0.1364 |
| • Head (%) | 13.0% | 9.0% | 7.0% | 0.8049 |
| • Neck (%) | 4.0% | 0.0% | 8.0% | 0.4519 |
| Tingling |  |  |  |  |
| • Feet (%) | 83.0% | 39.0% | 67.0% | **0.0006\*** |
| • Lower Legs (%) | 61.0% | 27.0% | 42.0% | 0.0675 |
| • Upper Legs (%) | 46.0% | 23.0% | 17.0% | **0.0037\*** |
| • Hands (%) | 70.0% | 32.0% | 46.0% | **0.0332\*** |
| • Lower Arms (%) | 43.0% | 14.0% | 20.0% | **0.0362\*** |
| • Upper Arms (%) | 43.0% | 14.0% | 12.0% | **0.0033\*** |
| • Torso (%) | 35.0% | 14.0% | 7.0% | **0.0046\*** |
| • Head (%) | 26.0% | 19.0% | 7.0% | 0.0500 |
| • Neck (%) | 13.0% | 9.0% | 8.0% | 0.8049 |

